# Benefits and Risks of Chloroquine and Hydroxychloroquine in The Treatment of Viral Diseases: A Meta-Analysis of Placebo Randomized Controlled Trials

**DOI:** 10.1101/2020.04.13.20064295

**Authors:** Jing Wang, Li Yu, Kefeng Li

**Affiliations:** Department of Critical Care Medicine, Yantai Yuhuangding Hospital, Yantai, Shandong 264000, China; Division of Pulmonary, Critical Care and Sleep Medicine, School of Medicine, University of California, San Diego, CA, 92037, United States; Department of Clinical Oncology, Shengjing Hospital of China Medical University, Shenyang, Liaoning 110004, China; Moores Cancer Center, School of Medicine, University of California, San Diego, CA, 92037, United States; Department of Medicine, School of Medicine, University of California, San Diego, CA, 92037, United States

## Abstract

**Background and Objective:** Recently, in the scramble to find drugs to treat COVID-19, chloroquine (CQ) and its derivative hydroxychloroquine (HCQ) have rapidly gained the public’s attention. In this study, we conducted a meta-analysis of randomized clinical trials (RCTs) to evaluate the efficacy and safety of CQ and HCQ in the treatment of viral diseases.

**Methods:** We searched PubMed, EMBASE, Cochrane Central, Web of Science, Clinical Trials Registries, CNKI, Wanfang Data, CQVIP, and Preprint Servers through April 4, 2020, for randomized controlled trials (RCTs) that examined the efficacy and safety of CQ and HCQ against viral infection. We analyzed pooled data on the overall efficacy, the relative risks over the placebo, and the prevalence of adverse events. Trial sequential analysis (TSA) was also performed to evaluate the random errors in the meta-analysis. Potential moderators of drug-placebo efficacy differences were analyzed by meta-regression.

**Results:** The analysis included 11 RCTs with 2613 adult patients. Both the plasma viral load (standard mean difference: 0.29, 95% CI: −1.19 - 1.76, *P* = 0.70) and the improvement of clinical symptoms (odds ratio: 2.36, 95% CI: 0.81 - 6.92, *P* = 0.11) were not different between the intervention and placebo arm. There was significant heterogeneity for the efficacy assessment, which was primarily explained by the mean patients’ age and the sample size. Compared to the placebo, CQ and HCQ had increased risk of mild adverse events (risk ratio: 1.51, 95% CI: 1.35 - 1.70, *P* < 0.05, TSA adjusted 95% CI: 1.31 - 2.19), which were statistically significant in nervous, integumentary, and gastrointestinal systems. The most common adverse events were observed in the nervous system, with the pooled prevalence of 31.4 % (95% CI: 10.5% - 56.7%).

**Conclusions:** Insufficient data were available to support the antiviral efficacy of CQ and HCQ due to the high heterogeneity caused by patients’ age. Mild side effects are expected for the current antiviral dose regimens of CQ and HCQ. Treatment outcomes may be enhanced by better-selected patients based on age and well-controlled adverse events.

This meta-analysis was registered on OSF (ID: https://osf.io/386aw)

## Introduction

Recently, the outbreak of COVID-19 has been declared as a pandemic by the World Health Organization (WHO). As of April 11, 2020, the coronavirus COVID-19 is affecting 185 countries and territories around the world, with 1,777,517 confirmed cases and over 108,862 deaths(1). Tremendous efforts have been made worldwide to search the effective antiviral drugs against COVID-19.

Chloroquine (CQ) and its derivative hydroxychloroquine (HCQ) are decades-old generic medicine used to treat malaria and rheumatoid conditions such as arthritis (2). In various studies, CQ had been demonstrated remarkable efficacy against emerging viral diseases such as HIV, severe acute respiratory syndrome (SARS), chikungunya, Zika virus, and influenza (3-7). The broad-acting antiviral activities and anti-inflammatory effects of CQ, obtained primarily from *in vitro* and animal studies, lead to the hypothesis that it may also be useful in the treatment of viral diseases. Randomized controlled trials (RCTs) are considered one of the best study designs to examine the efficacy and safety of an experimental drug. To date, a number of RCTs had been conducted to test the efficacy of CQ and HCQ against various viral infections (8). However, most of these trials had small sample sizes, and the results were inconsistent. Meta-analysis can overcome these limitations by increasing the sample size and, thus, statistical power to generate the best estimation. So far, no studies have performed either descriptive or quantitative meta-analysis of CQ and HCQ in the treatment of viral diseases.

Even though CQ is generally considered to be safe based on the safety data from anti-malarial treatment, the margin between the therapeutic and toxic dose is narrow(9). Additionally, weekly doses used in malaria might not be sufficient to reach the lower therapeutic window for its antiviral use(10).

CQ has been recommended in the Guidelines for the Treatment of Pneumonia Caused by COVID-19 issued by the National Health Commission of China, and the dose was adjusted later due to the potential adverse complications(11). Recently, the U.S. FDA has issued an emergency use authorization for hydroxychloroquine and chloroquine used in the clinical trials for COVID-19 patients. Therefore, understanding the previous evidence for benefit or harm from CQ/HCQ in the context of antiviral treatment is of immediate clinical importance.

In this study, we performed a systematic review and meta-analysis to evaluate the efficacy and adverse events of chloroquine and hydroxychloroquine in the treatment of viral diseases. We included all placebo-controlled randomized clinical trials (RCTs) containing CQ and HCQ as the antiviral therapy since its introduction in 1938 till April 3^rd^, 2020. We also performed univariate and multivariate meta-regression analyses to identify the moderators of drug-placebo differences. The results of this meta-analysis would help the design of clinical trials for the treatment of COVID-19 using CQ and HCQ.

## Materials and Methods

We performed the study in adherence to the Preferred Reporting Items for Systematic Reviews and Meta-Analysis (PRISMA) guideline (The checklist in the supplemental material).

### Search strategy

We performed a comprehensive literature search of articles through the following databases without date limitation: PubMed, EMBASE, The Cochrane Library, Web of Science, China National Knowledge Infrastructure (CNKI), Wanfang Data, and CQVIP. We also searched clinical trial registries, including ClinicalTrials.gov, Chinese Clinical Trial Registry, and EU Clinical Trials Register. The search was updated to April 4, 2020, and not restricted by language. Moreover, we contacted the corresponding authors of the included trials for any missing information. The search terms were presented in the supplemental material. The reference list was also checked for relevant articles.

### Inclusion and exclusion criteria

The PICOS inclusion criteria for selecting the studies for this meta-analysis were as follows: (1) Participants: patients with the viral infection and treated with chloroquine (CQ) or hydroxychloroquine (HCQ); (2) Interventions: CO or HCQ versus placebo; (3) Outcomes: primary outcomes included plasma viral load, the improvement of clinical symptoms, and adverse events associated with CQ (or HCQ) treatment; (4) Type of studies: In this meta-analysis, only randomized controlled trials (RCTs) were included. The exclusion criteria were as follows: 1) studies involving animals; 2) studies on non-viral diseases; 3) meeting abstracts, letters to the editor, case reports, and reviews.

### Data extraction and quality assessment

All potentially eligible articles were independently evaluated, and the information was extracted by two authors (J.W. and K.L.). Disagreements were resolved by discussion with a third person (L.Y.). For each study, the following items were extracted: first author, year of publication, country, type of viral infection, the total number of patients in the experimental or placebo group, mean age of the patients, drug dose, treatment duration, plasma viral load, number of patients with improved clinical symptoms, and the number and types of adverse events in both CQ/HCQ and the placebo arms. For the published RCTs only reporting the median, range and the size of the trial for plasma viral load, we used the following equation to estimate the mean and standard deviation without assuming the distribution of the underlying data (12):

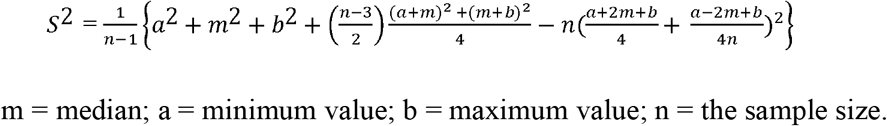

We used the Common Terminology of Clinical Adverse Events (CTCAE) categorization to identify grades 3 to 5 as severe and CTCAE grades 1 to 2 as mild. Missing data were requested directly from authors. Cochrane risk of bias tool in randomized trials (RoB 2.0) was used to assess the risk of bias for each of the included studies by two authors (J.W. and J.Z.) independently.

### Data synthesis and meta-analysis

The conventional meta-analysis was conducted using MedCalc 19.2.0 (MedCalc Software Ltd, Belgium). We used standard mean differences (SMDs) for the continuous outcome (plasma viral load) and odds ratios for the improvement of clinical symptoms based on the number of patients with improved symptoms on each arm and the sample size. The prevalence of adverse events in different organ systems was expressed as the proportion (%) and 95% confidence intervals (CIs) using the random-effects model. We used relative risk ratios and their 95% CIs for the assessment of adverse events compared to the placebo. Studies with zero-cell counts (No adverse events in one or both arms) were replaced with a fixed value of 0.5, according to Cochrane Handbook for Systematic Reviews of Interventions v5.1.0. We assessed heterogeneity using the I^2^ test and the Cochran Q statistic. If significant heterogeneity was not present (I^2^ < 50%), we used a fixed-effects model to pool outcomes. A random-effects model was used to calculate the pooled risk ratios and 95% CIs if significant heterogeneity was present (I^2^ > 50%). The publication bias was assessed qualitatively by the visual estimate of the funnel plots and quantitatively by the Begg’s and Egger’s tests. Sensitivity analyses were conducted by using an alternative effect measure (risk ratios vs. odd ratios), and statistical model regarding heterogeneity (random vs. fixed-effects).

### Trial sequential analysis

We performed trial sequential analysis (TSA) to explore whether cumulative data was appropriately powered. Trial sequential analysis (version 0.9.5.10, Copenhagen, Denmark) was used for cumulative meta-analysis(13). The type I error (α) and power (1 - β) were set as 5% and 80%, respectively. A 25% risk ratio reduction was used. The heterogeneity correction in the trial sequential analysis was set to variance-based, and the random-effects model was applied.

### Univariate and multivariate meta-regression analysis

We performed the meta-regression analyses to explore the potential moderators that may affect the efficacy of CQ. We conducted the univariate meta-regression analysis of publication year, mean age, trial site (country), virus types, sample size, daily dose, treatment duration, and cumulative dose. Moderators that were significant in univariable analyses were included in the multivariable meta-regression models. We also meta-regressed daily dose and treatment duration to explore the associations between these variables and the side effects. Both univariate and multivariate meta-regression analyses were conducted using Comprehensive Meta-Analysis 3.0 (Biostat Inc., NJ, USA). The regression coefficients (minimum estimate), 95% CIs, 2-sided *P* values, and Tau square were reported.

### Subgroup analysis

Subgroup analyses included stratification by the age of the patients to test the efficacy of CQ in two different groups (<38 vs. ≥38 years old, median line). We also performed the subgroup analyses to test the relative risk of CQ for different grades of adverse events (severe vs. mild) in various organ systems (nervous system, gastrointestinal system, integumentary system, and vision system.

## Results

### Literature search and flow diagram

The electronic searches identified 4314 records, and a further 65 records identified from clinical trial registration websites and other sources. Deleting duplicate references reduced this to 2482 records, of which 1897 were excluded based on examining the abstracts. Full-text articles were obtained for 33 publications. From these, 22 papers were excluded, leaving 11 placebo randomized controlled trials (RCTs) for final inclusion for quantitative synthesis and meta-analysis(14-24). The numbers identified at each stage through from initial searching to quantitative analyses, and the reasons for excluding studies, are given in a PRISMA 2009 flow diagram (Fig. 1).

**Fig. 1.**
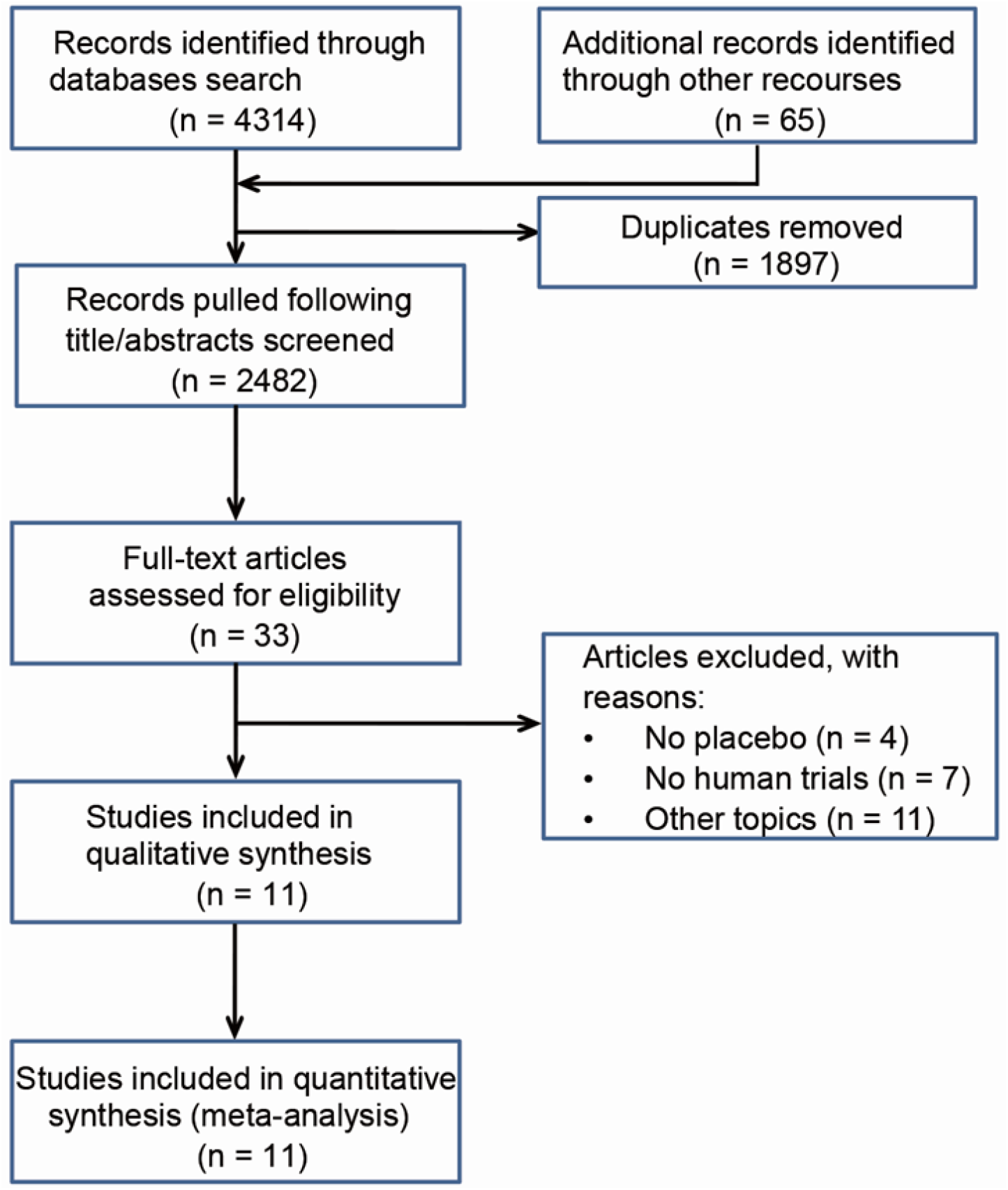
Flow chart of the included studies.

### Study characteristics

A total of 2613 patients were evaluated across the 11 RCTs. Table 1 shows the summary of the included trials, and Table S1 gives the details for adverse events in each organ system. Out of 11 RCTs, 2 studies were for the treatment of chikungunya virus, 2 studies were for Dengue virus, 1 for influenza, 1 for COVID-19, 2 for hepatitis, and 3 for HIV. The median dose of chloroquine base for the treatment of viral infection is 310 mg per day (IQR: 250 - 600 mg). The median duration of treatment was 56 days (IQR: 5 - 84 days). For all studies, the primary endpoint was the effectiveness of the treatment, with adverse events reported as secondary outcomes.

**Table 1.** Selected characteristics of the included studies.

| Author | Year | Country | Patients No. |  | Age<sup>*</sup> | Virus | Dose | Treatment Duration | Improved in CQ | Improved in Placebo | SMD of viral load in plasma (95% CI) | Adverse Events |  |
| --- | --- | --- | --- | --- | --- | --- | --- | --- | --- | --- | --- | --- | --- |
|  |  |  | CQ | Placebo |  |  |  |  |  |  |  | CQ | Placebo |
| Débora | 2019 | Brazil | 31 | 30 | 37.7 ± 16.1 | Hepatitis | 250 mg/day | 36 months | 20 | 13 | N/A | 17 | 5 |
| Pierre | 2018 | France | 19 | 27 | 40.3 ± 12.8 | Chikungunya | 600 mg D1-3, 300 mg D4-5 | 5 days | 8 | 6 | 2.41 (1.63 - 3.16) | N/A | N/A |
| Payam | 2016 | Iran | 6 | 4 | 49 (25 - 60) | Hepatitis | 150 mg/day | 8 weeks | 6 | 0 | -1.1 (-2.45 - 0.26) | 6 | 3 |
| Jeffrey | 2016 | USA | 35 | 35 | 46 (36 - 50) | HIV | 250 mg/day | 12 weeks | 0 | 0 | 0.31 (0.13 - 0.47) | N/A | N/A |
| Marcos | 2013 | Brazil | 63 | 66 | 32.7 ± 11.5 | Dengue | 600 mg/day | 3 days | 12 | 0 | N/A | 2 | 0 |
| Nicholas | 2011 | Singapore | 757 | 749 | 23.5 (22.1 - 31.6) | Influenza | 300mg D1-7, 300mg per week | 12 weeks | 712 | 730 | N/A | 341 | 249 |
| Vianney | 2010 | Viet Nam | 278 | 276 | 22 (18 - 27) | Dengue | 600 mg D1-2, 300 mg D3 | 3 days | 0 | 0 | N/A | 35 | 11 |
| Xavier | 2008 | France | 27 | 27 | 18 - 65 | Chikungunya | 600 mg D1-3, 300 mg D4-5 | 5 days | 0 | 0 | N/A | 14 | 0 |
| Chen | 2020 | China | 31<sup>#</sup> | 31 | 44.6 ± 16.1 | COVID-19 | 400 mg/day | 5 days | 25<sup>#</sup> | 17 | N/A | 2<sup>#</sup> | 0 |
| Nicholas | 2012 | UK | 42<sup>#</sup> | 41 | 37.1 ± 7.7 | HIV | 400 mg/day | 48 weeks | N/A<sup>#</sup> | N/A | 2.42 (1.33 - 4.31) | 41<sup>#</sup> | 26 |
| Kirk | 1995 | USA | 19<sup>#</sup> | 19 | 39.1 ± 6.6 | HIV | 800 mg/day | 8 weeks | N/A<sup>#</sup> | N/A | -0.71 (-1.37 - -0.056) | N/A<sup>#</sup> | N/A |

<sup>\*</sup>: mean and standard deviation or median with IQR or min-max range. <sup>#</sup>: HCQ. Additional characteristics were listed in Supplemental Table 1.

The results of the Cochrane risk of bias assessments for all studies are summarized in Fig. S1. Based on the information we collected, 1 had some concerns on the randomization process, 1 trial had the deviation of intended intervention for one patient. Three trails had some concerns about the selection of outcome reporting. All included RCTs were judged to be low risk in the overall risk of bias evaluation (Fig. S1).

### The efficacy of CQ and HCQ in the treatment of viral diseases

CQ and HCQ were reported to have direct antiviral activity mostly *in vitro* studies(3, 25, 26). In addition, these drugs showed anti-inflammatory and immunomodulatory effects in animal studies, which were proposed to be useful for the treatment of some clinical symptoms caused by viral infection(6, 27). We thus evaluated the efficacy of CQ and HCQ from the included RCTs using two outcomes: plasma viral load and clinical symptoms.

As shown in Fig. 2A, the plasma viral load in patients with CQ or HCQ was even slightly higher than that in the placebo group (SMD: 0.29, 95% CI: −1.18 - 1.76). Additionally, the meta-analysis demonstrated no significant difference in the improvement of clinical symptoms caused by viral infection between CQ (and HCQ) and the placebo (odds ratio: 2.36, 95% CI: 0.81 - 6.92, *P* > 0.05) (Fig. 2B). No significant publication bias was observed (Fig. S2), and the pooled results were consistent using the alternative statistical model and different measures in the sensitivity analysis (Fig. S3). We further conducted the trial sequential analysis (TSA) to determine the reliability of our meta-analysis (Fig. S4). The cumulative z curve fell within the futility zone (zone of indifference), and the TSA adjusted odds ratio was also insignificant between two groups (TSA adjusted odds ratio: 2.34, 95% CI: 0.73 - 7.47, *P* > 0.05).

**Fig. 2.**
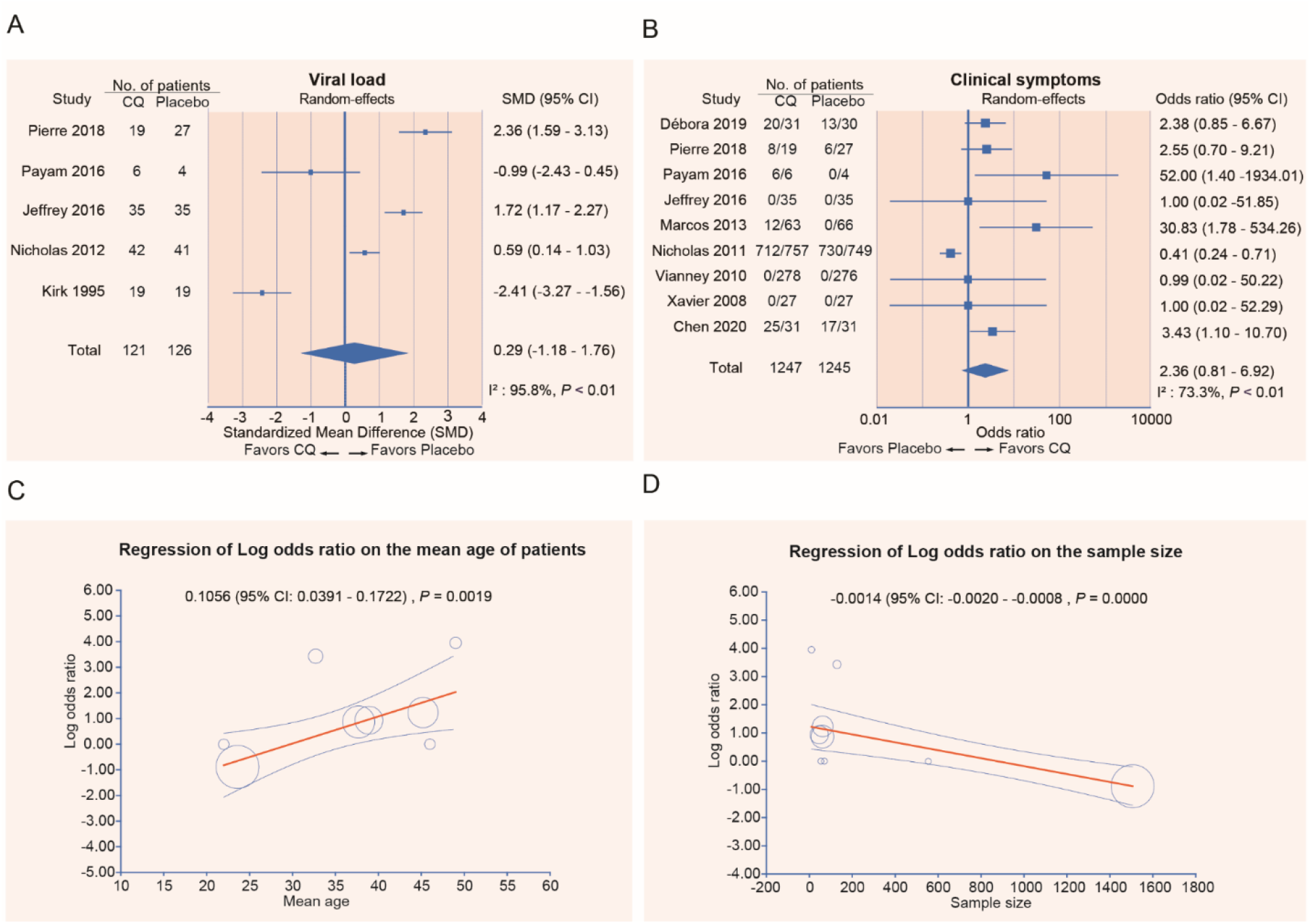
Plasma viral load and the improvement of clinical symptoms for patients taking CQ (or HCQ) and the placebo. (A) The standard mean differences (SMDs) of plasma viral load. (B) The odds ratio of the improvement of clinical symptoms between two arms. (C) Meta-regression analysis showed a significant association between the age of patients and the effect sizes. (D) The association between the sample sizes in the trails and the effect sizes. The coefficients (95% CI) and *P* values were listed.

We performed both univariate and multivariate meta-regression analysis to explain the significant heterogeneity of efficacy among the included studies (Table S2). In the univariate model, we found that mean age and the sample size are the significant moderators of effect sizes (Fig. 2C and 2D). Interestingly, an increase of effect size was observed with the increase of patients’ mean age ranging from 23.6 to 49 (coefficient: 0.1056, 95% CI: 0.039 - 0.172, *P* = 0.0019) (Fig. 2C). The strong association between the sample size and the efficacy of chloroquine was diminished after the adjustment of virus types in the multivariate meta-regression model (Table S2). Subgroup meta-analysis stratified by patients’ age (below and above the median of 38) showed that compared to the placebo, chloroquine significantly improved the clinical symptoms in patients with age over 38 (Odd ratio: 3.31, 95% CI: 1.54 - 7.12, *P* = 0.02), and such therapeutic effect was not observed in the younger group (22 - 37) (95% CI of the odds ratio: 0.33 - 9.91, *P* = 0.49) (Fig. S5). However, the cumulative z-curves in TSA did not cross the calculated trial sequential monitoring boundaries for benefits, indicating that the random errors might be present in the subgroup meta-analysis, and there is still no conclusive evidence of superiority for CQ and HCQ (Fig. S4B and S4C).

### The risk for all, mild and severe adverse events

Adverse events were reported in 35.0% (458/1308) of patients randomly assigned to chloroquine and in 22.5% (294/1305) of patients randomly assigned to the placebo (Table 1). The summary relative risk (RR) of adverse events was 1.52 (95% CI: 1.36 - 1.71), which indicated an increased risk of adverse events for CQ and HCQ over the placebo (Fig. 3A). Both the heterogeneity (I^2^ = 38.7%, *P* = 0.09) and the publication bias (Begg’s *P* = 0.94 and Egger’s *P* = 0.05) were insignificant (Fig. 3A and Fig. S6A). The pooled effects were not changed using odds ratios and alternative statistical methods in the sensitivity analysis (Fig. S7). Additionally, Trial sequential analysis supported the meta-analysis findings (Fig. S8). The cumulative z curve crossed the calculated trial sequential monitoring boundary, indicating that the information size was sufficient to find firm evidence.

**Fig. 3.**
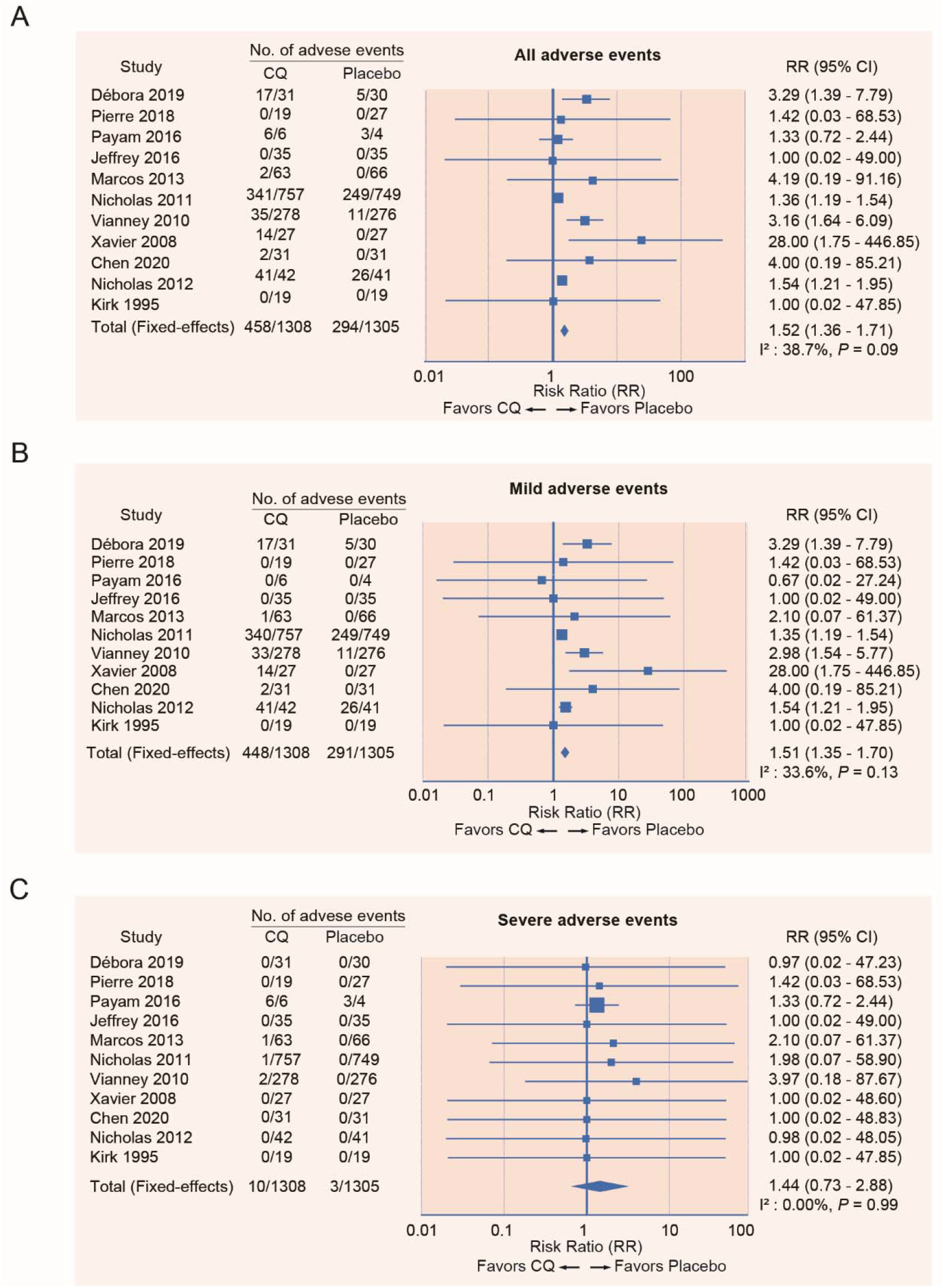
Risk of adverse events in patients with viral diseases who received CQ (or HCQ) versus the placebo from randomized clinical trials. (A) Forest plot of all adverse events in patients treated with CQ or HCQ versus controls. (B) Forest plot of mild adverse events. (C) Forest plot of severe adverse events.

We next grouped the patients based on the severity of adverse events using the common terminology of clinical adverse events (CTCAE). We found that patients treated with chloroquine for viral infection were more likely to experience mild adverse events (pooled RR: 1.51, 95% CI: 1.35 - 1.70) (Fig. 3B). In contrast, the pooled risk ratio of severe adverse events for chloroquine over the placebo was insignificant (pooled RR: 1.44, 95% CI: 0.73 - 2.88) (Fig. 3C). No evidence of publication bias was identified (Fig. S6B and S6C), and the meta-analysis results for both mild and severe adverse events were robust in sensitivity analyses (Fig. S9 and S10).

### Risk of adverse events in different organ systems

We next performed the meta-analysis of the prevalence of adverse events in different organ systems. In the intervention arms of these studies (n = 1308), the most common adverse events were observed in the nervous system (headache and dizziness) with the pooled prevalence of 31.4% (95% CI: 10.5% - 56.7%), followed by the integumentary system (rash, pruritus, skin darkening and thinning hair) of 20.9% (95% CI: 8.7% - 47.3%), gastrointestinal system (nausea, vomiting and diarrhea) of 14.5% (95% CI: 9.2% - 20.8%), vision (blurred vision and retinopathy) of 4.3% (95% CI: 1.7% - 8.1%) and heart (QT prolongation) of 0.08% (1 out of 1308) (Fig. 4 and Fig. S11).

**Fig. 4.**
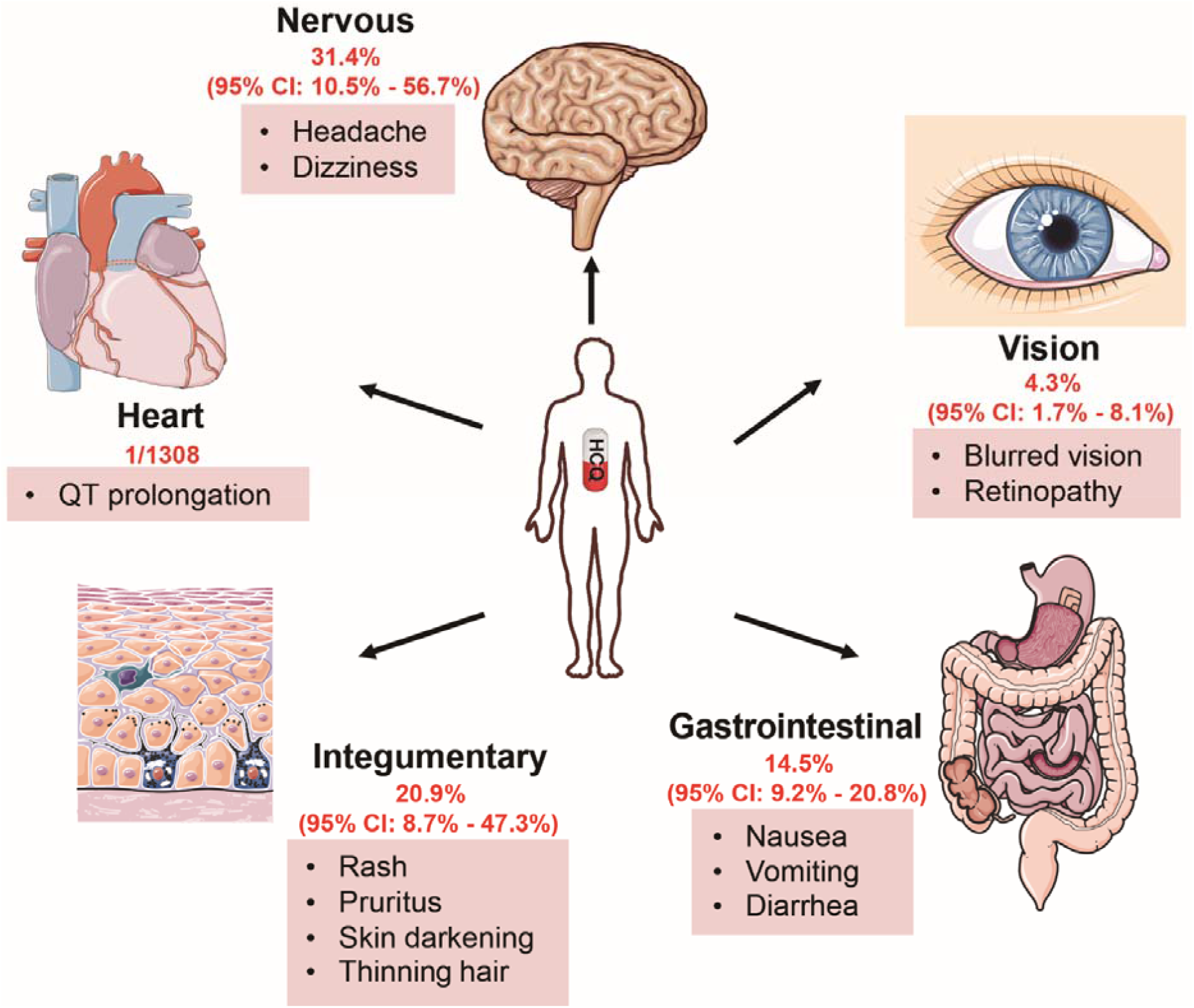
Meta-analysis of the prevalence of adverse events in the major human organ systems for CQ and HCQ. A random-effects model was used. N = 1308 patients in the intervention arm.

Compared with patients in the placebo arm, those treated with chloroquine were at higher risk for side effects in the nervous system (RR: 2.12, 95% CI: 1.30 - 3.45) (Fig. 5A), integumentary system (RR: 2.23, 95% CI: 1.45 - 3.43) (Fig. 5B), gastrointestinal system (RR: 2.52; 95% CI: 1.91 - 3.34) (Fig. 5C). The risk of adverse events in the vision system for patients with chloroquine was not significantly different from those with the placebo (RR: 2.08, 95% CI: 0.35 - 12.37) (Fig. 5D). This might be due to the low prevalence of adverse events in the vision system. No publication bias was detected (Fig. S12), and the sensitivity analysis using a different statistical model or effect measure did not significantly change the primary outcome (Fig. S13 - S16).

**Fig. 5.**
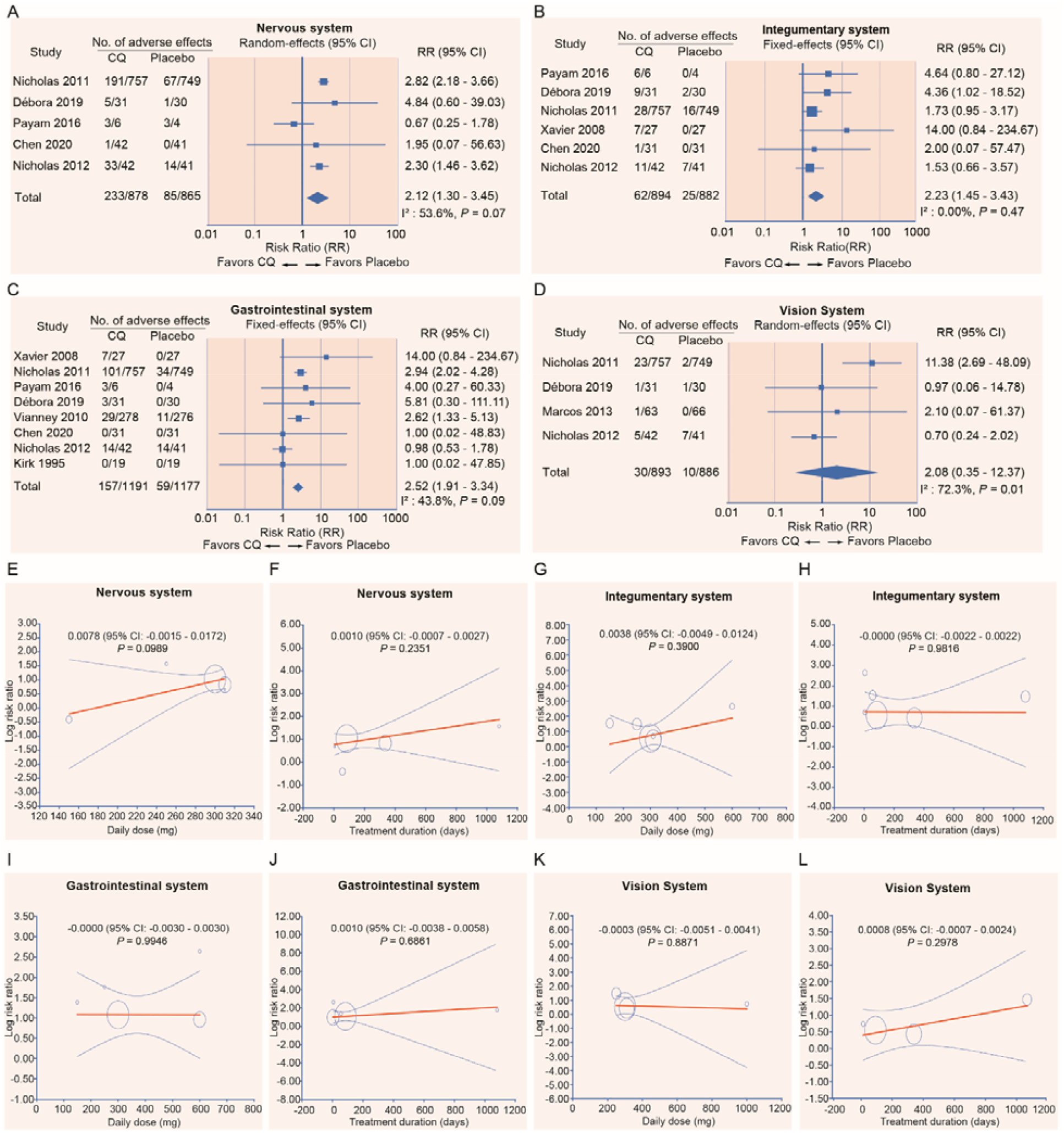
Risk of adverse events and the associations with daily dose and treatment duration of CQ (or HCQ) in different organ systems. (A - D) Forest plots of risk ratios of adverse events in the nervous system (A), integumentary system (B), gastrointestinal system (C), and vision system (D). (E - H) The associations of adverse events with daily dose and treatment duration in the nervous system (E and F), integumentary (G and H), gastrointestinal (I and J), and vision (K and L). Multivariate meta-regression analysis was performed using the random-effects model. The model was adjusted by sample size and mean age. The coefficients (95% CI) and *P* values were listed. The doses were converted to chloroquine base.

We further evaluated whether the increased risk of adverse events with chloroquine in different organs was associated with total daily dose and treatment duration. The multivariate meta-regression analysis did not identify significant correlations between the risk of side effects and daily dose (150 - 600 mg, chloroquine base, min - max) or treatment duration (3 - 1080 days, min - max) (Fig. 5E - 5L).

## Discussion

This meta-analysis of 11 RCTs (2613 patients) finds no evidence for the efficacy of chloroquine (CQ) or hydroxychloroquine (HCQ) in treating the viral diseases. However, there is firm evidence of an increased risk of mild adverse events in patients treated with CQ and HCQ compared to the placebo. No significant difference was identified concerning the severe adverse events between CQ (or HCQ) and the placebo. The pooled prevalence of side effects in different organ systems was as follows: nervous system (31.4%) > integumentary system (20.9%) > gastrointestinal system (14.5%) > vision (4.3%) > heart (0.08%). The risk of adverse events was not significantly increased with the increase of daily dose from 150 mg to 600 mg.

A conceptual strength of our analysis is that it was based on the randomized trials for chloroquine and hydroxychloroquine in the treatment of viral diseases, including 1 RCT trial for COVID-19. We also narrowed our analysis only to side effects and potential toxicities related to the dose regiments used for viral infection, which will be more informative for the ongoing and future clinical trials for COVID-19.

Another major strength of our analysis is that we included all available relevant data from completed studies of CQ and HCQ meeting our inclusion criteria. For the missing or ambiguous information, we contacted the authors and obtained the original data, if possible. This limits the risk of selective outcome reporting in our analysis.

Even though CQ and its hydroxy derivative HCQ have been demonstrated to have antiviral and anti-inflammatory activities mostly through in vitro and animal studies, they were not shown to be superior to the placebo in the improvement of clinical symptoms and viral clearance in our meta-analysis. Similarly, two recent pilot clinical trials in individuals with COVID-19 found no difference in the rate of virologic clearance between CQ treatment and controls without drug treatment or with standard treatment (Interferon α)(28, 29). However, our result had to be interpreted with caution due to the notable heterogeneity of the outcome (efficacy) across the included RCTs. Interestingly, we found that a great percentage of this heterogeneity can be explained by patients’ mean age and the sample size. Compared to the patients aged between 38 and 49, younger patients demonstrated less responses to CQ and HCQ treatment. It was reported that chloroquine is under-dosed for children(30). Our meta-analysis further highlighted that the doses need to be optimized for young patients for the treatment of viral diseases.

In our meta-analysis, we found that CQ and HCQ do not appear to increase the risk of severe adverse events over the placebo. It might be due to the low prevalence rate of severe events. Several individual cases of sudden cardiac death had been reported in COVID-19 patients treated with HCQ(31, 32). Therefore, the potential cardiotoxicity of CQ and HCQ, such as QT prolongation, has to be closely monitored during the clinical trial, especially for the combination use of HCQ and azithromycin(32).

Our analysis has some limitations. First, we did not conduct a subgroup analysis for the efficacy of CQ and HCQ on different types of viruses due to the small sample size in each study and the high heterogeneity among the included RCTs. Second, we assumed that the non-reporting of adverse event data was the result of no adverse event since the report of adverse events in RCTs had been required in the Consolidated Standards of Reporting Trials (CONSORT) extension for harms in 2004(33). If the adverse events were observed, but not reported, then there is the possibility that we may have overestimated the drugs’ safety. Third, adverse events in clinical trials are usually reported using the Common Terminology of Clinical Adverse Events (CTCAE). The process is highly subjective and relies on investigators’ recognition and identification of syndromes of interest.

## Conclusion

Both the public and clinicians must be aware that the value of chloroquine and hydroxychloroquine for the treatment of any viral diseases is not established. More large-scale, high-quality RCTs are still required to prove the efficacy of CQ and HCQ for COVID-19. The patients’ age is an important moderator that may affect the treatment outcome. Mild adverse events, most commonly in the nervous system, are expected, and potential severe cardiotoxicity has to be closely monitored during the treatment.

## Data Availability

All the data were presented in the manuscript and supplemental materials

## Authors’ contributions

J.W. and K.L. contributed to the conception of the study. K.L. contributed to the design of the study. JW and L.Y. contributed to the systematic review. J.W., and L.Y. designed the search strategy, screened the abstracts and full texts, acquired the data, and judged the risk of bias in the studies. JW, L.Y. and KL contributed to the data analysis. J.W. and K.L. contributed to the manuscript draft. JW, L.Y., and K.L. contributed to the manuscript revision. All authors read and approved the final manuscript.

## Acknowledgements

We would like to thank Dr. Jingbo Zhai from the Research Center of Tianjin University of Traditional Chinese Medicine (TCM) for his assistance on meta-analysis methodology.

## References

1. Dong E, Du H, Gardner L. An interactive web-based dashboard to track COVID-19 in real time. Lancet Infect Dis. 2020.

2. Schrezenmeier E, Dorner T. Mechanisms of action of hydroxychloroquine and chloroquine: implications for rheumatology. Nat Rev Rheumatol. 2020;16(3):155–66.

3. Vincent MJ, Bergeron E, Benjannet S, Erickson BR, Rollin PE, Ksiazek TG, et al. Chloroquine is a potent inhibitor of SARS coronavirus infection and spread. Virol J. 2005;2:69.

4. Savarino A, Cauda R, Cassone A. On the use of chloroquine for chikungunya. Lancet Infect Dis. 2007;7(10):633.

5. Savarino A, Di Trani L, Donatelli I, Cauda R, Cassone A. New insights into the antiviral effects of chloroquine. Lancet Infect Dis. 2006;6(2):67–9.

6. Savarino A, Boelaert JR, Cassone A, Majori G, Cauda R. Effects of chloroquine on viral infections: an old drug against today’s diseases? Lancet Infect Dis. 2003;3(11):722–7.

7. Fedson DS. Confronting an influenza pandemic with inexpensive generic agents: can it be done? Lancet Infect Dis. 2008;8(9):571–6.

8. Lu CC, Chen MY, Chang YL. Potential therapeutic agents against COVID-19: What we know so far. J Chin Med Assoc. 2020.

9. Touret F, de Lamballerie X. Of chloroquine and COVID-19. Antiviral Res. 2020;177:104762.

10. Olafuyi O, Badhan RKS. Dose Optimization of Chloroquine by Pharmacokinetic Modeling During Pregnancy for the Treatment of Zika Virus Infection. J Pharm Sci. 2019;108(1):661–73.

11. coronavirus McgoHCoGPfcitton. Expert consensus on chloroquine phosphate for the treatment of novel coronavirus pneumonia. Zhonghua Jie He He Hu Xi Za Zhi. 2020;43(3):185–8.

12. Hozo SP, Djulbegovic B, Hozo I. Estimating the mean and variance from the median, range, and the size of a sample. BMC Med Res Methodol. 2005;5:13.

13. Wetterslev J, Thorlund K, Brok J, Gluud C. Trial sequential analysis may establish when firm evidence is reached in cumulative meta-analysis. J Clin Epidemiol. 2008;61(1):64–75.

14. Raquel Benedita Terrabuio D, Augusto Diniz M, Teofilo de Moraes Falcao L, Luiza Vilar Guedes A, Akeme Nakano L, Silva Evangelista A, et al. Chloroquine Is Effective for Maintenance of Remission in Autoimmune Hepatitis: Controlled, Double-Blind, Randomized Trial. Hepatol Commun. 2019;3(1):116–28.

15. Roques P, Thiberville SD, Dupuis-Maguiraga L, Lum FM, Labadie K, Martinon F, et al. Paradoxical Effect of Chloroquine Treatment in Enhancing Chikungunya Virus Infection. Viruses. 2018;10(5).

16. Peymani P, Yeganeh B, Sabour S, Geramizadeh B, Fattahi MR, Keyvani H, et al. New use of an old drug: chloroquine reduces viral and ALT levels in HCV non-responders (a randomized, triple-blind, placebo-controlled pilot trial). Can J Physiol Pharmacol. 2016;94(6):613–9.

17. Jacobson JM, Bosinger SE, Kang M, Belaunzaran-Zamudio P, Matining RM, Wilson CC, et al. The Effect of Chloroquine on Immune Activation and Interferon Signatures Associated with HIV-AIDS Res Hum Retroviruses. 2016;32(7):636–47.

18. Borges MC, Castro LA, Fonseca BA. Chloroquine use improves dengue-related symptoms. Mem Inst Oswaldo Cruz. 2013;108(5):596–9.

19. Paton NI, Lee L, Xu Y, Ooi EE, Cheung YB, Archuleta S, et al. Chloroquine for influenza prevention: a randomised, double-blind, placebo controlled trial. Lancet Infect Dis. 2011;11(9):677–83.

20. Tricou V, Minh NN, Van TP, Lee SJ, Farrar J, Wills B, et al. A randomized controlled trial of chloroquine for the treatment of dengue in Vietnamese adults. PLoS Negl Trop Dis. 2010;4(8):e785.

21. De Lamballerie X, Boisson V, Reynier JC, Enault S, Charrel RN, Flahault A, et al. On chikungunya acute infection and chloroquine treatment. Vector Borne Zoonotic Dis. 2008;8(6):837–9.

22. Chen Z, Hu J, Zhang Z, Jiang S, Han S, Yan D, et al. Efficacy of hydroxychloroquine in patients with COVID-19: results of a randomized clinical trial. medRxiv. 2020:2020.03.22.20040758.

23. Paton NI, Goodall RL, Dunn DT, Franzen S, Collaco-Moraes Y, Gazzard BG, et al. Effects of hydroxychloroquine on immune activation and disease progression among HIV-infected patients not receiving antiretroviral therapy: a randomized controlled trial. JAMA. 2012;308(4):353–61.

24. Sperber K, Louie M, Kraus T, Proner J, Sapira E, Lin S, et al. Hydroxychloroquine treatment of patients with human immunodeficiency virus type 1. Clin Ther. 1995;17(4):622–36.

25. Liu J, Cao R, Xu M, Wang X, Zhang H, Hu H, et al. Hydroxychloroquine, a less toxic derivative of chloroquine, is effective in inhibiting SARS-CoV-2 infection in vitro. Cell Discov. 2020;6:16.

26. Wang M, Cao R, Zhang L, Yang X, Liu J, Xu M, et al. Remdesivir and chloroquine effectively inhibit the recently emerged novel coronavirus (2019-nCoV) in vitro. Cell Res. 2020;30(3):269–71.

27. Park TY, Jang Y, Kim W, Shin J, Toh HT, Kim CH, et al. Chloroquine modulates inflammatory autoimmune responses through Nurr1 in autoimmune diseases. Sci Rep. 2019;9(1):15559.

28. Molina JM, Delaugerre C, Goff JL, Mela-Lima B, Ponscarme D, Goldwirt L, et al. No Evidence of Rapid Antiviral Clearance or Clinical Benefit with the Combination of Hydroxychloroquine and Azithromycin in Patients with Severe COVID-19 Infection. Med Mal Infect. 2020.

29. Chen J, Liu D, Liu L, Liu P, Xu Q, Xia L, et al. A pilot study of hydroxychloroquine in treatment of patients with common coronavirus disease-19 (COVID-19). J Zhejiang Univ (Med Sci). 2020;49(1):0.

30. Commons RJ, Simpson JA, Thriemer K, Humphreys GS, Abreha T, Alemu SG, et al. The effect of chloroquine dose and primaquine on Plasmodium vivax recurrence: a WorldWide Antimalarial Resistance Network systematic review and individual patient pooled meta-analysis. Lancet Infect Dis. 2018;18(9):1025–34.

31. Yazdany J, Kim AHJ. Use of Hydroxychloroquine and Chloroquine During the COVID-19 Pandemic: What Every Clinician Should Know. Ann Intern Med. 2020.

32. John R. Giudicessi, Peter A. Noseworthy, Paul A. Friedman, Ackerman MJ. Urgent guidance for navigating and circumventing the QTc prolonging and torsadogenic potential of possible pharmacotherapies for COVID-19. Mayo Clin Proc. 2020;95.

33. Ioannidis JP, Evans SJ, Gotzsche PC, O’Neill RT, Altman DG, Schulz K, et al. Better reporting of harms in randomized trials: an extension of the CONSORT statement. Ann Intern Med. 2004;141(10):781–8.

